# Dietary Exposure to Oxidized Lipids in US Infants

**DOI:** 10.1101/2022.06.22.22276754

**Authors:** Lisaura Maldonado-Pereira, Carlo Barnaba, Ilce Gabriela Medina-Meza

## Abstract

Baby Foods (BFs) and Infant formulas (IFs) are the main sources of nutrition for an infant throughout the 1^st^ year of life. A variety of enriched products are commercially available for parents seeking to fulfill their baby’s nutritional needs. Consequently, different bioactive lipids are present in BFs and IFs including dietary oxidative substances (DOxS) whose known toxicity has been associated with mutagenicity, cancer, and other chronic diseases. In this work, we performed an exposure assessment of 25 bioactive lipids on a total of 63 baby food samples (50 – IFs and 13 – BFs) commercially available in the US. To determine the dietary exposure to DOxS, we used EPA’s SHEDS-HT probabilistic model. Even though β-Sitosterol was the most exposed bioactive lipid with 75,4 µg/day, cholesterol was the most absorbed compound during the entire first year (19,320 µg/day). Additionally, we found 7α-hydroxycholesterol (7α-OH) as a potential DOxS biomarker of the BFs manufacturing process. This is the first time that an exposure assessment including DOxS ingested by infants after BFs and IFs consumption is performed, enabling much-needed information regarding these hazardous compounds and their potential effects on infants’ health.

## 1. Introduction

Diet, during the first year of life of a newborn, plays a crucial role to ensure an infant’s proper growth and brain development (Babawale et al., 2018; CDC, 2021b; Harris & Pomeranz, 2020; Timby, Domellöf, Hernell, Lönnerdal, & Domellöf, 2014). Despite the U. S. Dietary Guidelines for Americans recommendation for breastfeeding during the first 6 months, reports show that only 25% of babies are exclusively breastfed in the US (CDC, 2021a), and globally, only 40% have been estimated (Collective, 2017; Timby et al., 2021). Different circumstances influence the mother’s decision to rely mainly on infant formulations (IFs) instead of breast milk. After the baby is 6 months old, baby foods (BFs), also known as complementary foods, are commonly introduced to the infant, and it is not until the infant’s 1^st^ or 2^nd^ year that they will be eating almost the same foods as any other adult (CDC, 2021b).

Cholesterol content in human milk is typically comprised of 0.34 to 1.3 wt% based on total lipids (Jensen, 1999). On the contrary, standard IF doesn’t contain cholesterol but it is high in plant-based oils such as palm and soybean oil (Zubin Maslov, Hill, Lüscher, & Narula, 2021) which provide other lipid classes such as phytosterols and tocopherols also needed for a healthy growth and development processes. During weaning (4-6 months), BFs are gradually introduced. BFs consist of unsweetened, unsalted, no added starch or artificial flavors/colors puree of fruits, vegetables, grains, and blends of meats or poultry. All these ingredients are good sources of protein, iron, zinc, and other much-needed nutrients (Gerber, 2021a, 2021b).

The main purpose of IFs and BFs manufacturing relies on providing the best, most nutritious, and safest food for the most vulnerable population, infants. Both IF and BFs undergo industrial processing to guarantee their microbiological safety. Processing, in particular thermal processing, act on macro and micronutrients of raw material, potentially favoring changes in the nutritional properties of these foods. For instance, several lipid oxidation molecules, such as dietary oxysterols (DOxS), are formed because of food’s over-processing during their confection, storage, and even transportation processes.

The oxidation of cholesterol results in the accumulation of DOxS. The formation of DOxS is triggered by high temperature, hydrostatic pressure used for nonthermal pasteurization, light exposure, and aging of food (Cais-Sokolińska, Walkowiak-Tomczak, & Rudzińska, 2022; Chudy & Teichert, 2021; Lisaura Maldonado-Pereira, 2021; Risso et al., 2022; Sabolová et al., 2017). Several studies have summarized the biological activities of these compounds which have been extensively associated with different chronic diseases including pro-inflammatory and pro-oxidant activities, as well as promote carcinogenesis (Garcia-Llatas, Mercatante, López-García, & Rodríguez-Estrada, 2021; L. Maldonado-Pereira, Schweiss, Barnaba, & Medina-Meza, 2018; Poli et al., 2022; Willinger, 2019; Zmysłowski & Szterk, 2019). Our group recently studied the metabolomic signatures of liquid and powder infant formulations and proposed that 7-keto could be considered a biomarker of powder IFs (A. Kilvington, Barnaba, Rajasekaran, Laurens Leimanis, & Medina-Meza, 2021). Additionally, Risso and coworkers pointed out that the shelf-life is the primary determinant of DOxS after analyzing whole milk powders (Risso et al., 2022). Their study serves as another example of how DOxS could be used as markers to further quantify and characterize the nutritional quality and freshness, not only of ingredients but also of composite products such as commercial chocolate milk and whole powder milk. These findings highlight the potential relationship of DOxS loads with specific unit operations used in food processing. This knowledge may be a critical factor in identifying processing steps that may increase their formation in foods and could prevent infants’ exposure to these compounds and their long-term effect on their health. Previously, we assessed the dietary exposure of DOxS on infant formula-fed infants in the first 6 months of life (Alice Kilvington, Barnaba, Rajasekaran, Laurens, & Gabriela Medina-Meza, 2021). To date, the estimated exposure level of DOxS in infants (0-1 years old) has not been addressed.

The present study provides a dietary assessment of sterols and their oxidized derivatives using a probabilistic exposure assessment model during the first year of infant development for several sterols species (previously analyzed in our laboratory) present in various IFs and BFs commercially available in the US market (A. Kilvington et al., 2021; A. Kilvington, Maldonado□Pereira, Torres□Palacios, & Medina□Meza, 2019). The dietary exposure assessment will provide an estimation of the exposure extent of these compounds in infants which is the first step in conducting a risk assessment of these sterols and their oxidative species that could become a potential health risk in the infant’s community (Wei et al., 2022).

## 2. Methods

### 2.1. Stochastic Human Exposure and Dose Simulation (SHEDS-HT)

SHEDS-HT was coded in the R language (v. 2.15.3). Several modifications were performed to the code’s input files to enable the dietary exposure analysis of bioactive lipids including sterols (both animal and plant origin), squalene, and tocopherols. A complete description of the SHEDS-HT model can be found in Isaacs’ work (Isaacs et al., 2014). The following sections will explain the modifications employed in this analysis.

#### 2.1.1. Chemical Pathway and Scenario

A list of the compounds analyzed in this study is shown in **Table 1**. The chemical and physical properties of the compounds required by SHEDS-HT were obtained from The Metabolomics Innovation Centre and the ChemSpider webpages (Centre, 2021; ChemSpider, 2021). Concentration values of these chemicals in Infant Formulas (IFs) were obtained from previous work performed in the Food and Health Engineering Laboratory (FHEL) at Michigan State University (A. Kilvington et al., 2019).

**Table 1:** Chemical properties used in SHEDS-HT to evaluate exposure of the compounds of interest.

| Compound | Water Solubility (mg/L) | Absorption factor ($f_{abs}$) |
| --- | --- | --- |
| 7keto | 0.0027<sup>1</sup> | 0.12<sup>3</sup> |
| Triol | 0.000156<sup>1</sup> | 0.15<sup>3</sup> |
| 5,6alpha-Epoxy | 0.005104<sup>2</sup> | 0.28<sup>3</sup> |
| 5,6beta-Epoxy | 0.005104<sup>2</sup> | 0.32<sup>3</sup> |
| 7beta-OH | 0.48<sup>1</sup> | 0.42<sup>3</sup> |
| 7alpha-OH | 0.48<sup>1</sup> | 0.30<sup>3</sup> |
| 20alpha-OH | 0.57<sup>1</sup> | 0.40* |
| 22-OH | 0.65<sup>1</sup> | 0.40<sup>4</sup> |
| 24-OH | 0.40<sup>1</sup> | 0.40* |
| 25-OH | 0.24<sup>1</sup> | 0.40* |
| Campesterol | 0.05803<sup>5</sup> | 0.1431<sup>5</sup> |
| Beta-sitosterol | 0.02739<sup>5</sup> | 0.05074<sup>5</sup> |
| Stigmasterol | 0.0000112<sup>5</sup> | 0.040<sup>5</sup> |
| Brassicasterol | 0.06420<sup>5</sup> | 0.3121<sup>5</sup> |
| Alpha-Tocopherol | 0.003317<sup>5</sup> | 0.03394<sup>5</sup> |
| Beta-Tocopherol | 0.02826<sup>5</sup> | 0.03423<sup>5</sup> |
| Delta-Tocopherol | 0.2477<sup>5</sup> | 0.03477<sup>5</sup> |
| Gamma-Tocopherol | 0.02852<sup>5</sup> | 0.03424<sup>5</sup> |
| Cholesterol | 0.077038<sup>5</sup> | 0.314705<sup>5</sup> |
| Squalene | 0.004897<sup>5</sup> | 0.986721<sup>5</sup> |
| Desmosterol | 0.049427<sup>5</sup> | 0.378243<sup>5</sup> |

<sup>1</sup>The Metabolomics Innovation Centre (https://www.metabolomicscentre.ca/)
<sup>2</sup>ChemSpider – Search and share chemistry (www.chemspider.com)
<sup>3</sup>Value obtained from the paper: Osada, Sasaki, & Sugano, 1994
<sup>4</sup>Value obtained from the paper: Tranheim Kase et al., 2012
<sup>5</sup>Properties provided by SHEDS-HT
\*Absorption value of 22-OH was used for these compounds since no estimated value could be found for these specific DOxS (20alpha-OH, 24-OH, and 25-OH)

DOxS concentrations from Baby Foods (BFs) were obtained experimentally as is explained in sections 2.2-2.4. Since this study was focused on ingestion of DOxS because of BFs and IFs consumption, SHEDS-HT only needs to consider the dietary pathway and the direct ingestion scenario of the samples tested. In this study, the dietary exposures are calculated by determining the total daily mass of chemical intake for each simulated person via different foods. Concentration distributions for each food are provided as input. For each individual, daily concentrations (μg of sterol per g of food) are sampled (one for each food item). Dietary exposures are calculated as the sum (over food groups) of the food item’s concentration and mass of food consumed (as determined by the assigned food diary for the person). Exposure equation used by SHEDS-HT:

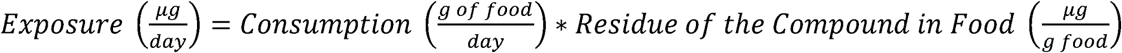

Intake dose estimates are calculated using distributions of route-specific fractional absorptions as it is shown below:

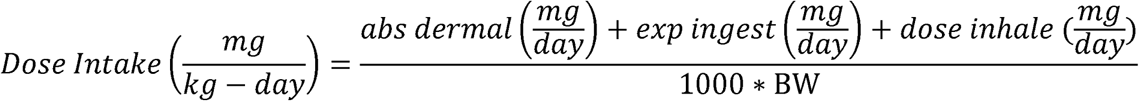

Where *abs dermal* if the absorption of the compound through the skin, *exp ingest* is the exposure of the compound through ingestion of the food item, and *dose inhale* is the dose of the compound inhaled by the infant.

Absorption values are obtained using the estimated absorption factor (f_abs_) of each bioactive lipid in the following equation:

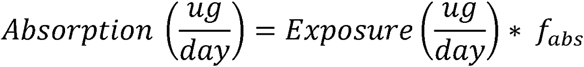

A full description of the exposure equations used in SHEDS-HT and their corresponding input parameter distributions are provided in Isaacs’ work (Isaacs et al., 2014).

#### 2.1.2. Population Module

Input-based data from the U.S. Census was used to generate a simulated population representative of the U.S. population in terms of age and gender. A population of 10,000 individuals was selected for this study. Afterward, Monte Carlo methods were used to assign relevant exposure factors and cohort-matched activity and food intake diaries to each person.

Food diaries were based on the National Health and Nutrition Examination Survey-What We Eat in America (NHANES-WWEIA) 1999−2018 two-day food intake diaries (USDA, 2018). Food diaries calculated the mass of each food group consumed by the individual.

### 2.2. Bioactive Lipids Extraction in Baby Foods

Bioactive lipids were extracted from 13 baby foods according to a slightly modified Folch’s (Folch, Lees, & Sloane Stanley, 1957) cold extraction method as is described in Maldonado-Pereira et al. (2022). Thirty grams of sample were minced and placed in a 500 ml glass bottle with a screwcap with 200 mL of chloroform:methanol solution (1:1, v/v). The sample was mixed for 15 min at 300 rpm. Homogenization was performed using an Ultra-Tourrax for 3 minutes. The bottle was kept in an oven at 60°C for 20 min before adding 100 mL chloroform. Then, the bottle was vortexed for 2 min following by filtration. The filtrate was mixed thoroughly with 100 mL of 1 M KCl solution. Samples were left overnight at 4 °C. Then, the lower phase containing lipids was collected and dried at 60°C with a vacuum evaporator at 25 in Hg. Total fat content was determined gravimetrically (L. Maldonado-Pereira, Barnaba, De los Campos, & Medina-Meza, 2021).

### 2.3. Determination of Total Cholesterol, Tocopherols, and Phytosterols Content in Baby Foods by GC-MS

A simultaneous quantification method for cholesterol, tocopherols, and phytosterols developed by our group was used (A. Kilvington et al., 2021). Cold saponification at room temperature was performed following Bonoli’s protocol with some modifications (Matteo Bonoli et al., 2008). Two hundred mg of lipid were mixed with 50 µg of 19-hydroxycholesterol and 140 µg of 5α-cholestane as internal standards for the determination of dietary COPs and total cholesterol, respectively. Subsequently, the sample was mixed with 10 mL of 1 N KOH solution in methanol and left under gentle agitation, and covered from the light for at least 15 h (Sander, Addis, Park, & Smith, 1989). One-tenth of the unsaponifiable matter was subjected to silylation. The sample was mixed with 100 µL of pyridine and 100 µL of the Sweeley’s reactive mixture (pyridine/hexamethyldisilazane/trimethylchlorosilane, 10:2:1, v/v/v) at 75°C for 45 min, dried under nitrogen stream and dissolved in 1 mL of n-hexane. One microliter of the silylated solution was injected into a gas chromatograph using a fitted with a Zebron™ZB-5HT (Phenomenex, Torrance, CA) capillary column (30 m × 0.25 mm × 0.25 μm). Oven temperature conditions were set up as follows: 260 to 300°C at 2.5°C/min, then from 300 to 320°C at 8°C/min, and finally hold at 320°C for 1 min. The injector and detector were both set at 320°C. Helium was used as the carrier gas at a flow rate of 54.0 mL/min, the split ratio at 1:50, and the pressure constant at 134 kPa.

### 2.4. DOxS profiling in Baby Foods

The other nine-tenths of the sample was used for the quantification of dietary COPs. Purification by NH_2_-SPE was performed according to the literature (Rose-Sallin, 1995). The enriched fraction was recovered and evaporated to dryness under a nitrogen stream. Subsequently, the purified fraction was silylated with Sweeley’s reactive mixture (at 75 °C for 45 min), dried under a nitrogen stream and dissolved in 1 mL of n-hexane.

Dietary DOxS were identify using the selected ion molecule (SIM) method with a GCMS-QP2010 SE single quadrupole Shimadzu Corporation (Kyoto, Japan). One μL of the silylated DOxS was injected into a single quadrupole (GCMS-QP2010 SE) with the following conditions: from 250 to 280 °C at 2°C/min, hold at 280 °C for 7 min, and from 280 to 315 °C at 1.5 °C/min. Helium was used as a carrier gas (flow rate of 0.37 mL/min); the split ratio was 1:15 and the pressure was 49.2 kPa. The interface temperature for the GC-MS was 320 °C, with the electron multiplier voltage set at 0 kV and the injector at 320 °C. A fused silica column (30 m x 0.25 mm i.d. x 0.25 um thickness) coated with 5% phenyl polysiloxane (Phenomenex Zebron ZB-5) was used.

### 2.5. Statistical Analysis

Descriptive statistics were obtained from the SHEDS-HT model. Mean, standard deviation, and the following quantiles: 5%, 25%, 50%, 75%, 90%, and 95%, were reported. Since the data did not follow a normal distribution after a Shapiro-Wilk test a non-parametric Matt-Whitney *U-* test was performed at a *p* < 0.05 significance level. Data normalization was performed for exposure, absorption, and dose intake results for comparison purposes. A sparse PLS-DA (sPLS-DA) was used to evaluate relationships between a single ‘omics data set. Additionally, a biomarker analysis, including the ROC curves, were performed using MetaboAnalyst in its web server version (Chong et al., 2018) All the statistical analysis were computed using RStudio (version 1.4.1717 © 2009-2021 RStudio, PBC)

## 3. Results and discussion

### 3.1 Bioactive Lipids Profiling in BF

A complete list of the BF samples, their codes, and results can be found in the Supplemental Material – **Table 1S and Table 2S**. Cholesterol was the most abundant compound in BF (**Figure 1S**), reaching a concentration value of 1,247.2 mg/100g fat for BF1 (**Table 1S**). A higher cholesterol value compared to the one reported in the nutritional facts label was observed for samples: BF1, BF6, and BF11. The items presenting higher fat content had mainly beef, turkey, and dairy products as the main ingredient in their preparation process. Plain meat BFs after the first 6 months of age is recommended by the CDC to avoid iron deficiency (CDC, 1998; Fox, Pac, Devaney, & Jankowski, 2004; Miller, 2013). Even though a higher fat content was observed for those specific BFs, fat consumption is normally promoted during the infants’ first years of life because of its essential role during the infants’ growth phase. Restricting fat before the age of two is not recommended and could be harmful during the infants’ growing stage (Allen & Myers, 2006; Hardy & Kleinman, 1994; Johnson, 1998; Öhlund, Hörnell, Lind, & Hernell, 2008).

Phytosterols comprised 39% of the total bioactive lipid’s distribution, followed by DOxS with only 2%. Tocopherols, desmosterol, and squalene were not detected in our BF samples.

β-Sitosterol was the dominant phytosterol which agrees with previous studies (Guadalupe García-Llatas et al., 2008; G. García-Llatas & Rodríguez-Estrada, 2011; A. Kilvington et al., 2021). The presence of high amounts of β-Sitosterol reflect the use of vegetable oils in the preparation of most BFs.

### 3.2 Dietary Exposure from IFs and BFs during Infant’s 1^st^ Year of Life

The equation used by SHEDS-HT to calculate the exposure value of each compound is shown above. Exposure results are reported in units of µg/day (**Table 3S**) and are based on the quantities of IFs and BFs ingested by infants between 0 and 1 year old reported by the NHANES-WWEIA 1999-2018. Overall, 59% of the most exposed bioactive lipids were phytosterols (**Figure 1**) - specifically, β-Sitosterol with 75,410 ug/day (**Figure 2**) - which is the most abundant group and lipid reported in our previous study (A. Kilvington et al., 2021). Cholesterol and campesterol were the 2^nd^ and 3^rd^ most exposed bioactive lipids. Among DOxS, 5,6β-epoxycholesterol, 7β-OH, and 7α-OH obtained the 8^th^, 9^th^, and 10^th^ highest exposure levels with 1,244.4, 191.8, and 165.5 µg/day, respectively. These DOxS have been related to several adverse effects on human health in previous studies such as cardiovascular diseases - atherosclerosis, hypertension, ischemic stroke-, neurological diseases, viral infections, and cancer (A. Kilvington et al., 2021; Lisaura Maldonado-Pereira, 2021; L. Maldonado-Pereira et al., 2018; Zmysłowski & Szterk, 2019), which arise concerns related to their consumption by infants.

**Figure 1:**
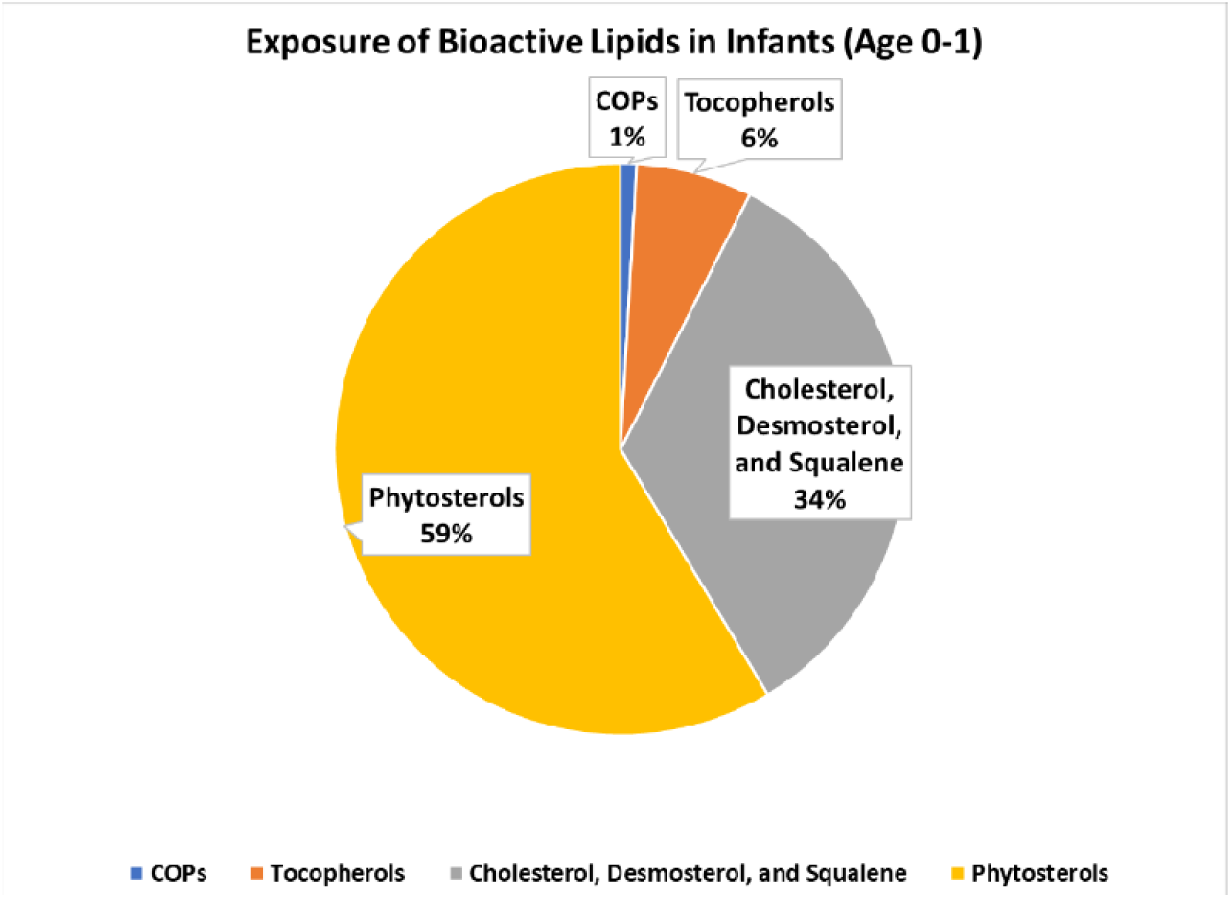
Bioactive Lipids Exposure in IFs and BFs Consumption during 1^st^ Year of Life

**Figure 2:**
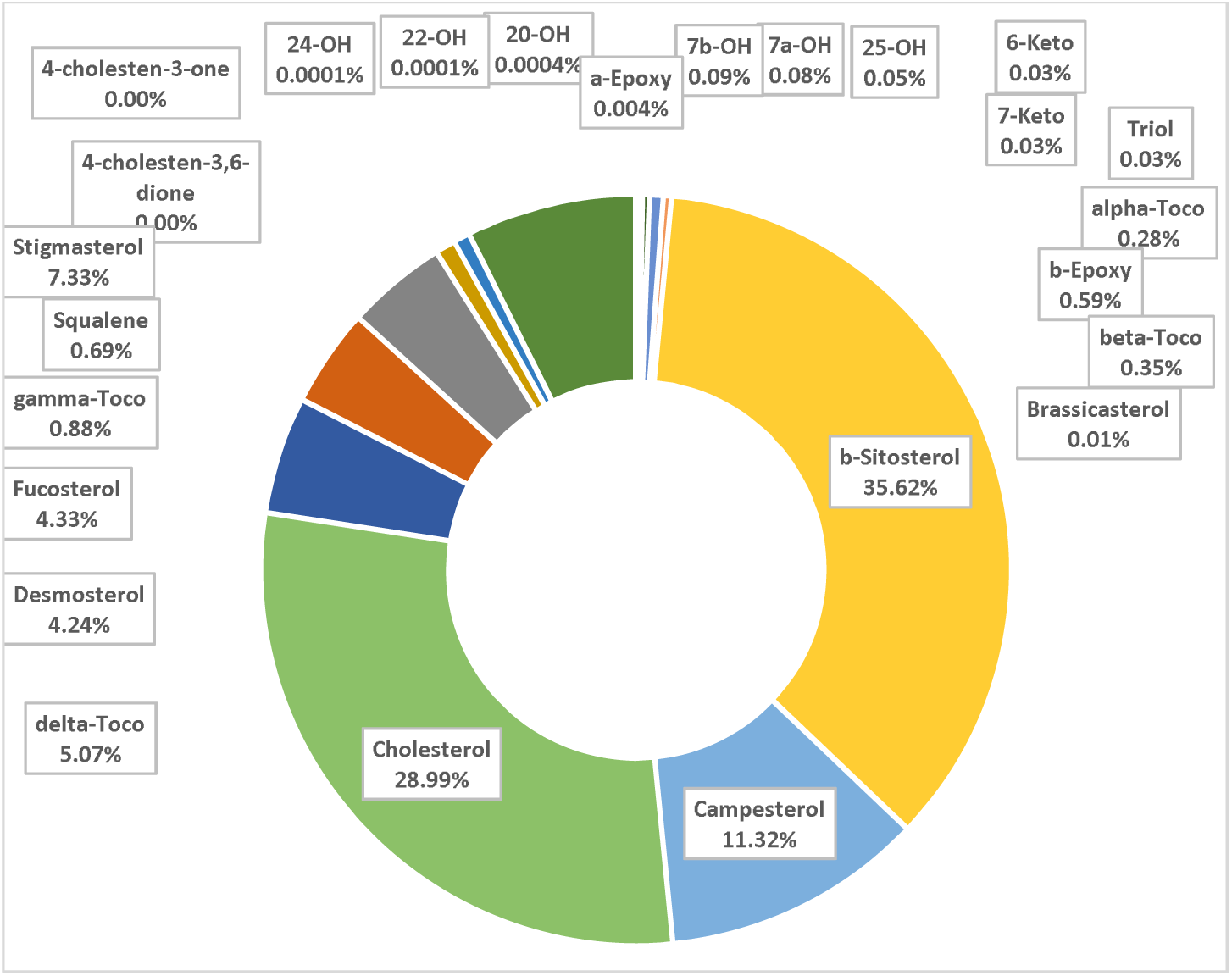
Percentages of the Bioactive Lipids Exposure Values in IFs and BFs after their Consumption during the 1st Year of Life according to Supplemental Material - Table 3S

### 3.1 Dietary Dose Intake and Absorption of Bioactive Lipids by Infants (0-1 years old)

The dietary dose intake of phytosterols (**Figure 3B**) was higher than any other bioactive lipid group which agrees with previous studies (Claumarchirant, Matencio, Sanchez-Siles, Alegría, & Lagarda, 2015; Hamdan, Claumarchirant, Garcia-Llatas, Alegría, & Lagarda, 2017).

**Figure 3:**
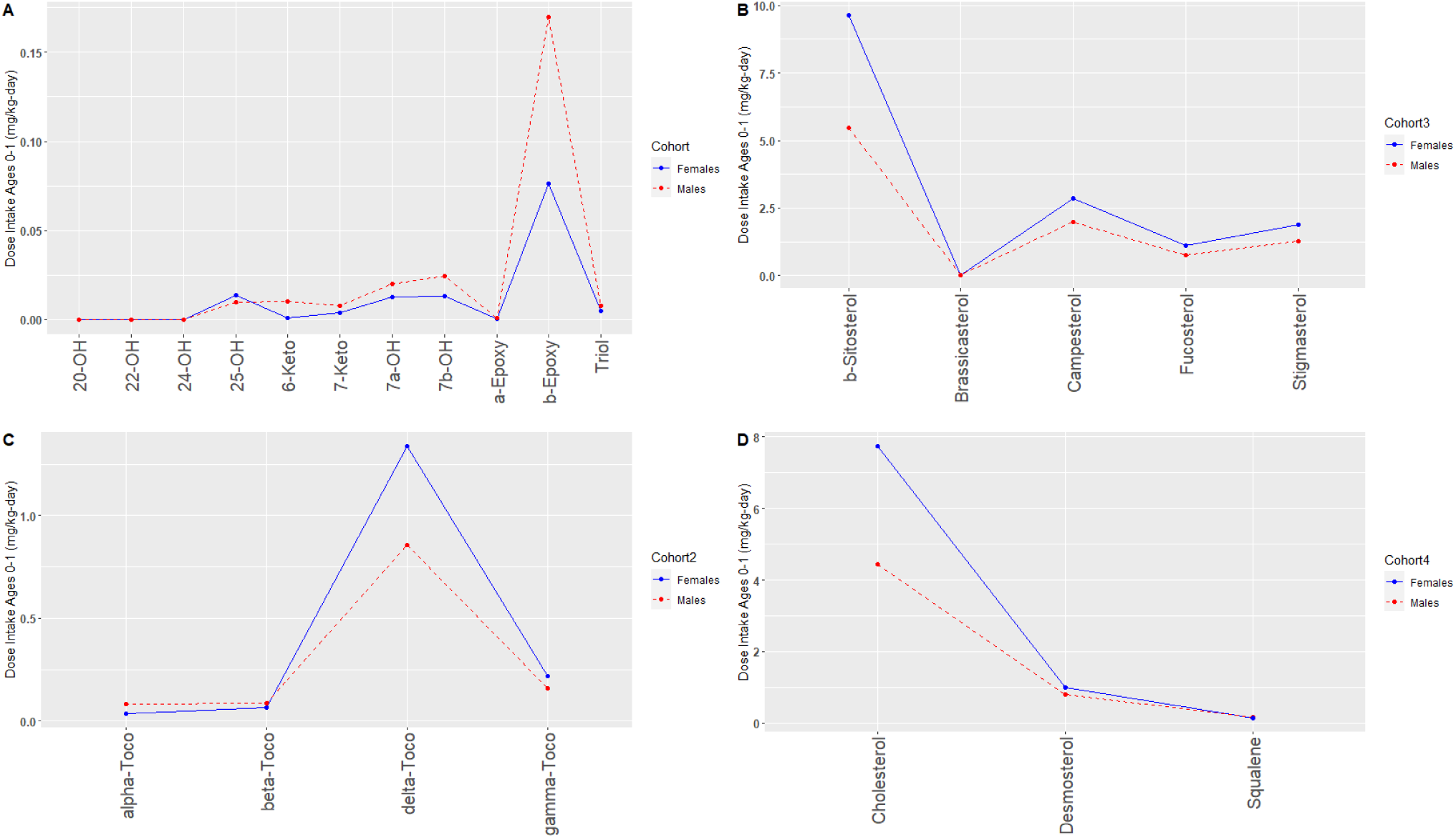
Dose Intake (mg/kg-day) of A) DOxS, B) Phytosterols, C) Tocopherols, and D) Cholesterol, Desmosterol, and Squalene

β-Sitosterol showed the largest dose intake among all bioactive lipids reaching a value of 9.64 mg/kg day in females (Hamdan, Sanchez-Siles, Garcia-Llatas, & Lagarda, 2018; Hamdan, Sanchez-Siles, Matencio, et al., 2018).

Interestingly, dietary dose intakes (mg/kg-day) were higher in male infants compared to females for most of the DOxS (5,6β-epoxycholesterol, 6-ketocholesterol, 7-ketocholesterol, 7α-OH, and 7β-OH) and α-tocopherol (**Figure 3A and 3C**). On the other hand, dietary dose intake of β-sitosterol, cholesterol, δ-tocopherol, and 25-OH were slightly higher for female infants.

Even though this difference in dose intake values between sexes was not significant, it highlights the effect of body weight’s average variability between females and males (body weight usually varies by only 0.79 kg) (CDC, 2001). In our previous study focused only on IFs consumption, the difference in dietary dose intake values between males and females was also not statistically significant (A. Kilvington et al., 2021).

The estimated absorption factors (f_abs_) provided a predicted absorption value (mg/kg-day) for each specific bioactive lipid analyzed in this model (**Figure 4A and 4B**). Cholesterol showed the highest absorption value with 1.90 mg/kg-day. Subsequently, β-sitosterol, campesterol, and desmosterol showed values of 0.38, 0.34, and 0.33 mg/kg-day, respectively.

**Figure 4:**
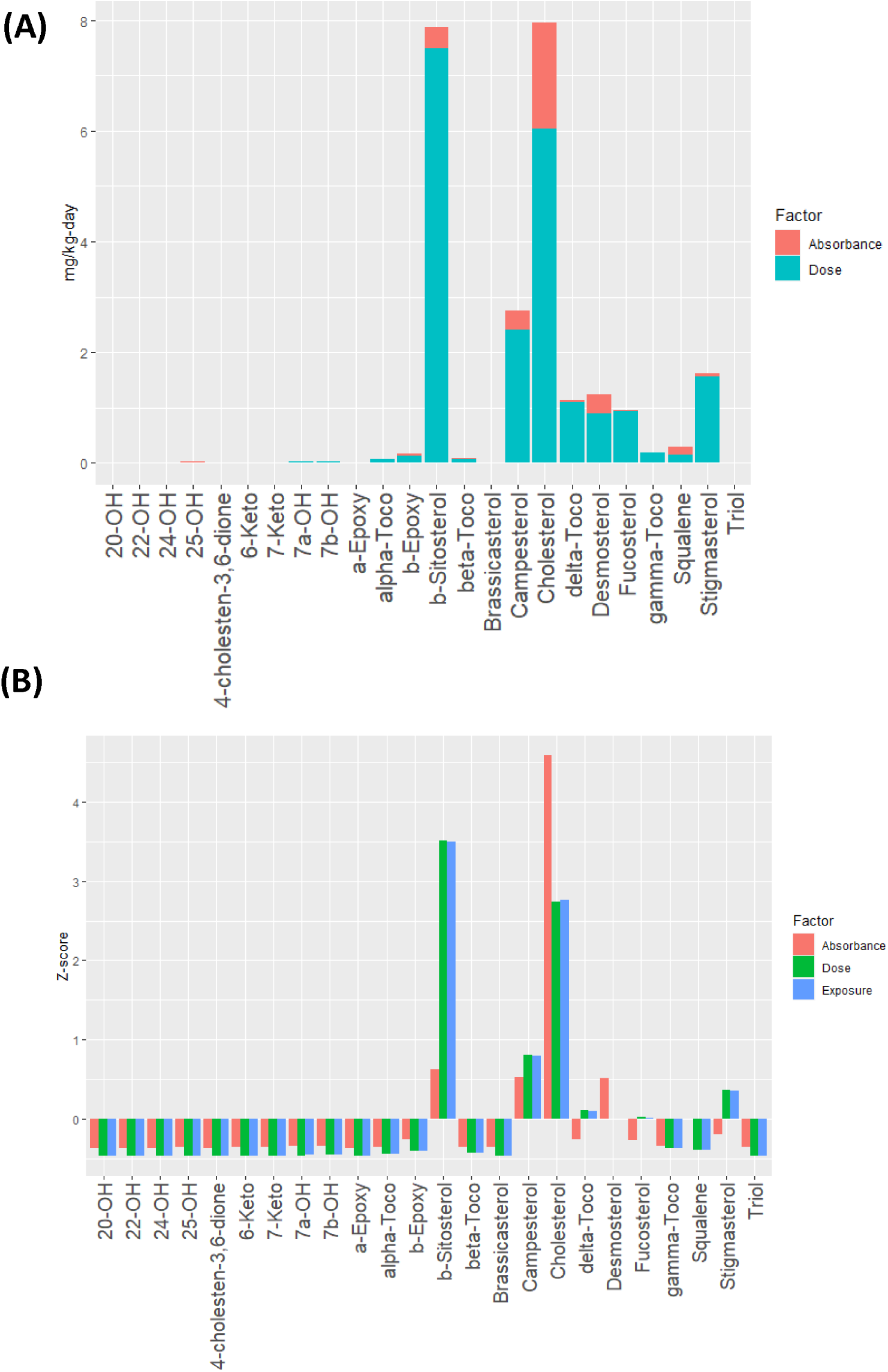
A) Comparison of absorption and Dose Intake. B) Normalization of Exposure, Dose Intake, and Absorption

Even though β-sitosterol was the compound with the highest exposure value, cholesterol was absorbed by 80% more. The bioaccessibility of β-sitosterol has been reported to be lower than other sterols, which explains the reason for being less absorbed than other sterols, despite its higher exposure level, and dietary dose intake (Claumarchirant et al., 2015).

All DOxS together showed an absorption value of 0.062 mg/kg-day, being 5,6β-epoxycholesterol the DOxS with the highest absorption value (0.040 mg/kg-day). There’s a clear distinction between DOxS absorption comparing 0-6 and 6-12 months of life of an infant. The presence of 7-keto in IFs was previously seen in metabolomics studies such as the one performed by our group (A. Kilvington et al., 2021), where 7-keto was proposed as a processing biomarker for powder IFs. Here, it is observed that it is not the most exposed, ingested, and absorbed sterol (**Figure 4A**) once solids foods are introduced into the infant’s diet. Therefore, it could be stated that it’s highly absorbed by infants, probably during just the first 6 months of life based on their dietary intake through ***only*** IFs consumption. With the introduction of solid foods past the first 6 months of life, 5,6β-epoxycholesterol resulted to be more absorbed. Absorption of these two DOxS raises concerns since their toxicity has been reported in previous studies in concentrations as low as 30.0 and 39.7 ng/mg of biological tissue, specifically in atherosclerosis patients (Micheletta et al., 2004). Maldonado-Pereira and coworkers provided a comprehensive review that reported the concentration of DOxS that exerted biological activities (L. Maldonado-Pereira et al., 2018). It is crucial to study in-depth the relationship between infants’ exposure and absorption of these DOxS with the future development of different chronic diseases.

### 3.2 Comparison of Bioactive Lipids Exposure, Dose Intake, and Absorption

Normalized values of exposure, dose intake, and absorption for all compounds are shown in **Figure 4B**. Cholesterol’s normalized absorption value was almost doubled compared to either its normalized exposure or dose intake values. It is known that massive amounts of cholesterol are needed for an infant’s growth (especially during the first 0-24 months of life) since every membrane requires cholesterol with especially high amounts in neuronal cells (Lawrence & Lawrence, 2022; Woollett & Heubi, 2000; Zubin Maslov et al., 2021). Therefore, such a high absorption rate of cholesterol was already expected.

Overall, 5,6β-epoxycholesterol was the most absorbed DOxS. It is noteworthy to mention that DOxS were absorbed in a significantly less proportion than cholesterol (**Figure 5**) as a result of their lower solubility in the mixed micelles and/or lower esterification inside the enterocytes (Garcia-Llatas et al., 2021). Additionally, the dominant presence of phytosterols in BF and IF could be exerting an antioxidant effect that promotes such high cholesterol absorption rate while it hinders further absorption of COPs. This high presence of phytosterols could result in a positive attribute of IF and BF by reducing the absorption of the DOxs and promoting cholesterol absorption which is much needed by infants at this early stage of life.

**Figure 5:**
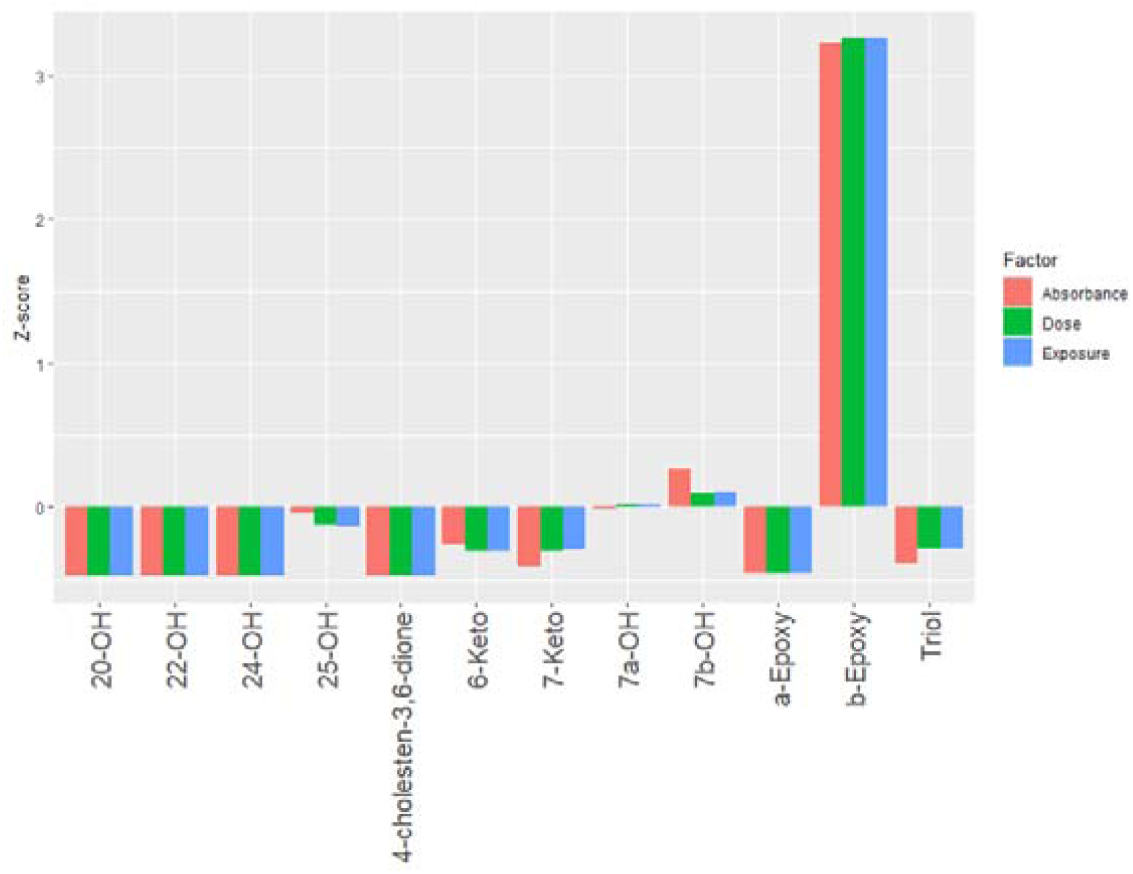
Normalization of Exposure, Dose Intake, and Absorption Only for COPs during 1^st^ Year of Life

For β-sitosterol, a higher dose intake was obtained after comparing the exposure level. As it was mentioned above, β-sitosterol is an abundant phytosterol in vegetable oils which is the main ingredient of IF and certain BFs. Additionally, it could be the exerting an antioxidant effect during the bioactive lipids absorption process after BFs and IFs consumption.

Lastly, since the absorption values for 25-OH, triol, and 7-keto were estimated values obtained from 2 different scientific sources (Centre, 2021; ChemSpider, 2021), an Absorption, Digestion, Metabolism, and Excretion (ADME) study is needed to obtained realistic absorption rates that will facilitate more accurate exposure assessments.

### 3.3 Dietary Dose Intake Comparison between SHEDS-HT and Theoretical Equation

After comparing the results from this exposure assessment with our previous study (Kilvington et al., 2021), a better idea of the dietary intake and potential absorption values was obtained. Kilvington and her co-worker estimated dose intake values using the theoretical equation:

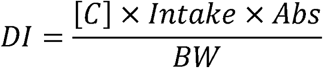

This equation was modified to analyze the ingestion and dose intake during the first 6 months of life of several bioactive lipids. Our previous work identified 7-keto as a potential biomarker for IFs. Lately, Risso and coworkers also suggested 7-keto as a biomarker in their study using powder milk samples (Risso et al., 2022).

Our results -which are derived from the equation previously mentioned used by SHEDS-HT-revealed that compared to the first 6 months of an infant where their diet is mainly comprised of IFs, 5,6β-epoxycholesterol, 7β-OH, and 7α-OH were the most absorbed DOxS from BFs consumption. 5,6β-epoxycholesterol, 7β-OH, and 7α-OH have been previously found in several types of meat and poultry samples (Chiu, 2018; Gehring et al., 2020; Grau, Codony, Grimpa, Baucells, & Guardiola, 2001; Khan et al., 2015; Musto, Faraone, Cellini, & Musto, 2014; Osada, Hoshina, Nakamura, & Sugano, 2000; Pie, 1991; Rant et al., 2019). Compared to IFs, BFs’ protein content commonly comes from different meat and poultry ingredients used during their confection. Interestingly, based on unpublished results from our lab, 5,6β-epoxycholesterol has been found present in human placenta samples. This finding could somehow relate the higher absorption rate of this DOxS with prenatal factors such as hormones present during the infant’s fetal development (Prins et al., 2022; Rodríguez-Rodríguez et al., 2018; Wu, Tian, & Lin, 2015)

Biomarker analysis for our BF and IF data revealed that a 7α-OH could be a DOxS processing biomarker for BFs (**Figure 6**). From these results and considering our previous study, we suggest that 7α-OH is a biomarker for BFs while 7-keto serves as a biomarker for powder IF. It is worth noting that 7-OH isomers are direct precursors of 7-keto. Differences in the type of thermal processing of each type of food (IF and BF) are the main reason that leads to the formation of certain DOxS, which suggests them as potential biomarkers of either BF or powder IF. This hypothesis is strongly supported by previous studies that have reported differences in DOxS dominance on the same food matrices cooked under different conditions (Francesc Guardiola, 2002; F. Guardiola, Bou, Boatella, & Codony, 2004; Lisaura Maldonado-Pereira, 2021; L. Maldonado-Pereira et al., 2018).

**Figure 6:**
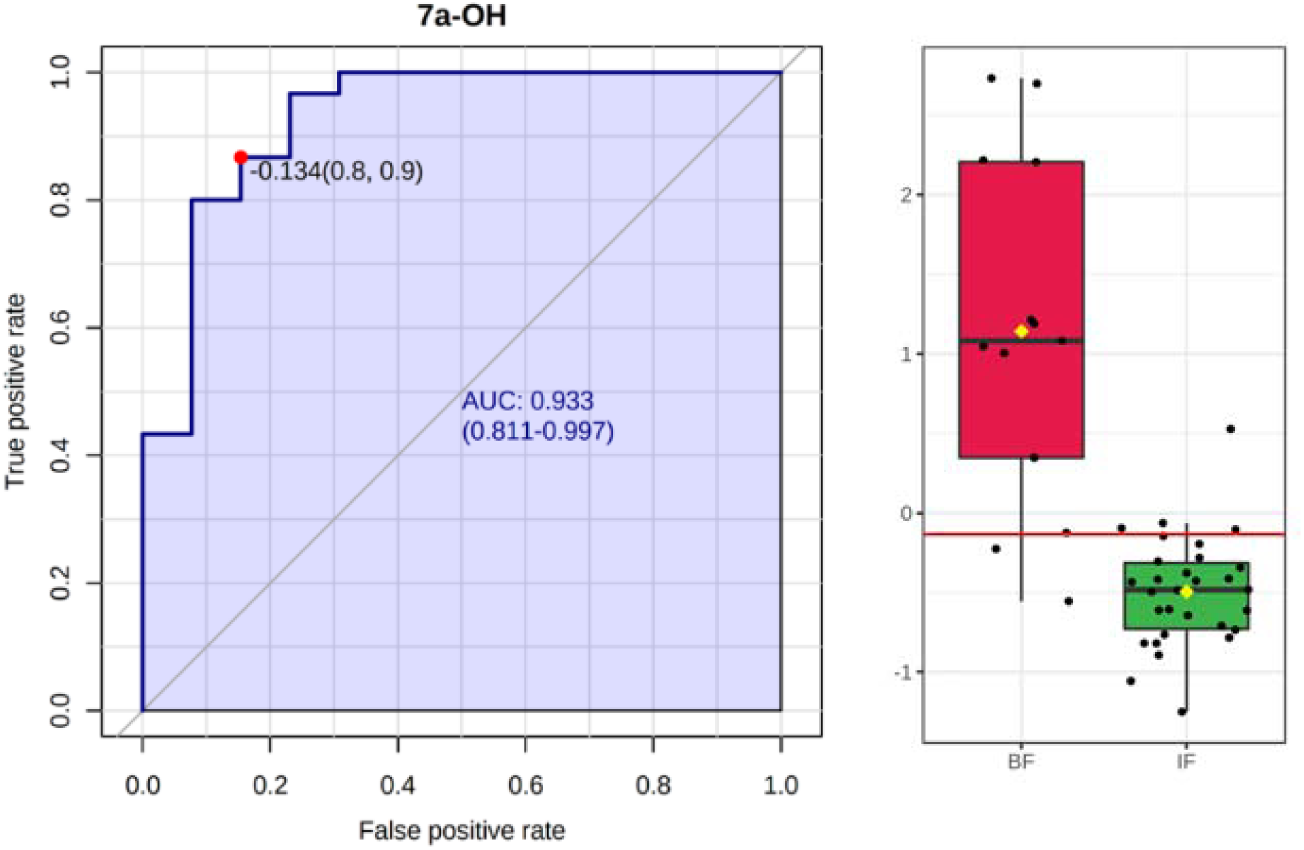
7α-OH as a Potential Biomarkers of BFs

An sPLS-DA was performed to evaluate the accuracy of our models. Data were classified by BF, Liquid IFs (L-IFs), and Powder IF (P-IFs) to highlight any potential differences in IF processing techniques. **Figures 7A** show that there is indeed a noticeable difference between the confection process of BF, L, and P that affects the values of the DOxS present in the samples.

**Figure 7:**
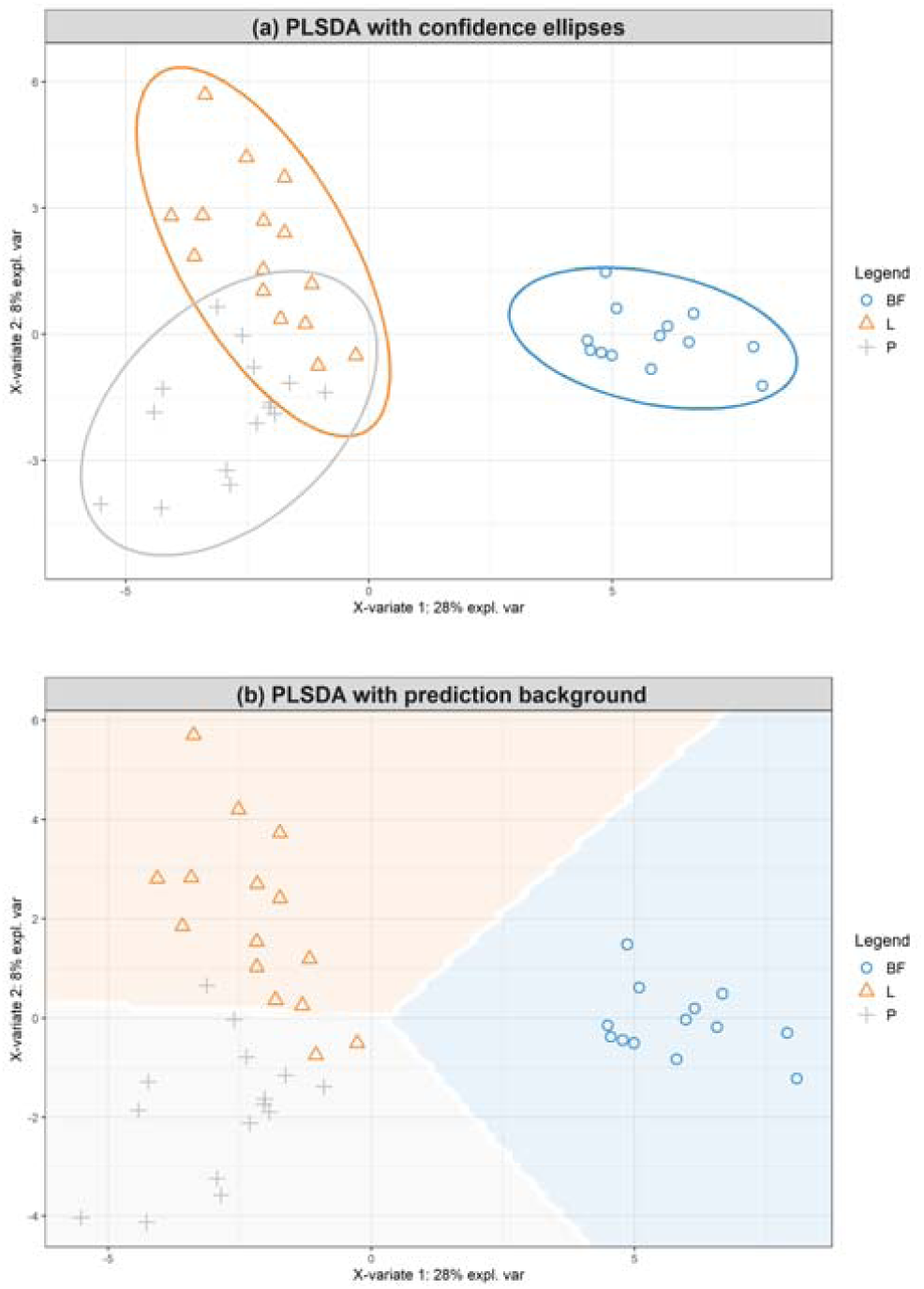
A) IF and BF data with confidence ellipses and B) Prediction Zones

Additionally, **Figure 7B** shows the prediction zone for other samples based on our data. In our model, only 3 out of 63 samples were incorrectly classified, reaching a 95.2% accuracy. Both of these figures provide intuition on how novel samples would be classified according to th generated model. Lastly, **Figure 8** shows a ROC for all 3 models (BF, L-IFs, and P-IFs) with highly positive sensitivity rates for each one (BF: 0.1, P-IFs: 0.9905, and L-IFs: 0.981, allowing a good prediction rate for future data.

**Figure 8:**
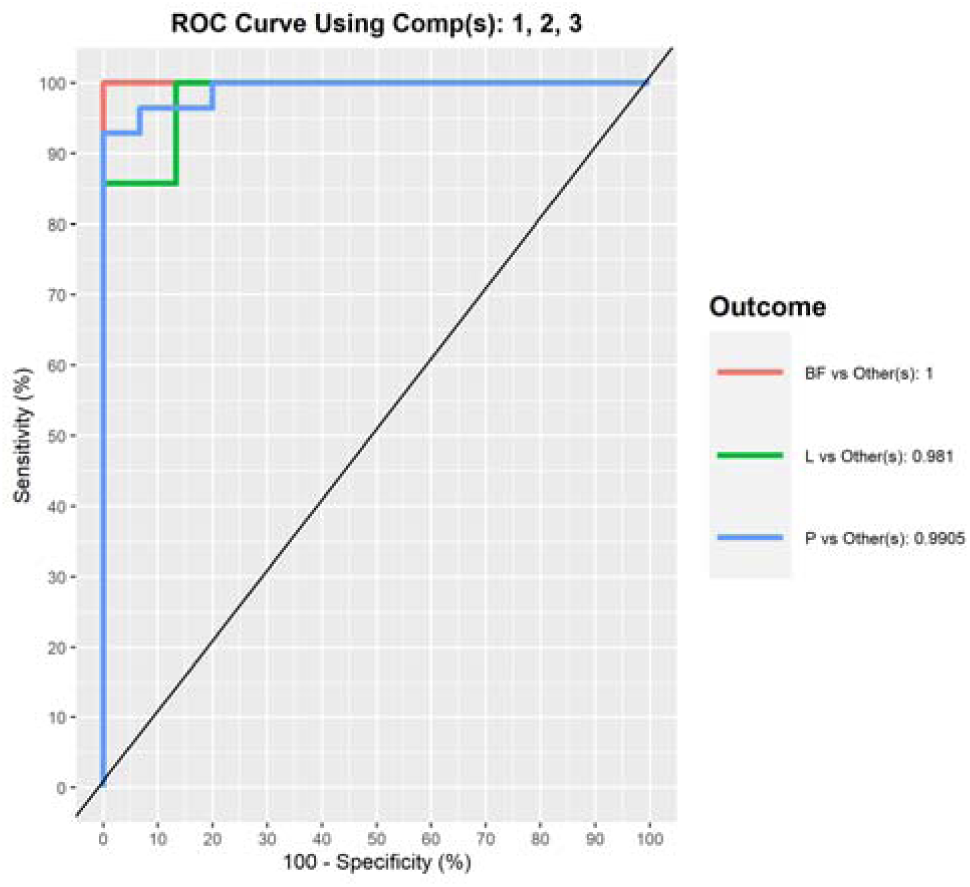
ROC for BF, Liquid Formula, and Infant Formula Models

Besides the use of 7α-OH and 7-keto as biomarkers of thermal processing techniques employed during the confection of BFs and IFs, it is important to keep in mind their biological toxicity that could become a potential hazard among infants. Comparing both DOxS, 7α-OH and 7-keto have been found in patients suffering different chronical diseases such as hypertension, multiple sclerosis, cancer, atherosclerosis, among others (L. Maldonado-Pereira et al., 2018). The lowest concentrations exerting a toxicological effect previously reported for 7α-OH and 7-keto were 0.004 and 0.006 µg/mL of blood plasma, respectively, in atherosclerosis patients. Since there is so little available information regarding the relationship between these compounds and the development of these diseases as a result of the consumption of IF s and BFs by infants, this vulnerable population could be potentially at a health risk. More accurate information regarding DOxS absorption values, their metabolism, and excretion rates is needed to start reducing this knowledge gap. Therefore, we suggest the development of an Absorption, Digestion, Metabolism, and Excretion (ADME) study followed by the completion of a risk assessment to identify the real hazard and protect our infants.

## 4. Conclusions

Bioactive lipid distribution of BF is dominated by cholesterol (59%) from animal based foods, followed by phytosterols (∼39%) from plant based foods. β-Sitosterol was the most abundant phytosterol as a result of its dominant presence in vegetable oils, which is the BF’s main ingredient.

Dietary exposure assesment showed that phytosterols had the highest exposure percentages (59%) compared to DOxS, tocopherols, cholesterol, squalene, and desmosterol. β-Sitosterol was the most exposed compound (0.38 mg/kg-day) but the 2^nd^ most absorbed compound because of its lower bioaccessibility which has been previously reported (Claumarchirant et al., 2015). Cholesterol was the most absorbed lipid with a 1.90 mg/kg-day.

5,6β-epoxycholesterol was the DOxS with the highest absorption value (0.040 mg/kg-day) which places it overall in the 7^th^ position among all bioactive lipids absorbed. Even though absorbance of all DOxS together accounted for only 1.86% (0.062 mg/kg-day), there’s evidence of their biological effect which raises concerns related to their consumption by infants. There is a critical need to deeper investigate the relationship of infants diet – especially during the 1 year of life – with future development of chronic diseases.

One of our model’s limitations includes the lack of information regarding the absorption fractions (f_abs_) of 25-OH, triol, and 7-keto. For the purpose of our study, these values were estimated from the available data found from 2 different scientific sources (Centre, 2021; ChemSpider, 2021). This is why the FHEL suggests and is currently conducting an Absorption, Digestion, Metabolism, and Excretion (ADME) study – especifically of DOxS - to obtain more accurate absorption values to enable a more precise exposure assessment. Despite this weakness, this is the first time that an exposure assessment including DOxS ingested by infants after BFs and IFs consumption is performed, enabling information regarding these hazardous compounds and their potential future effects on infants’ health.

Based on the biomarker analysis, 7α-OH could be a potential biomarker of BFs which could not be predicted by just analyzing the exposure and absorption data obtained from SHEDS-HT.

The difference in DOxS distribution between BF, IFs-L, and IFs-P as a result of processing techniques variability, was confirmed after an sPLS-DA was performed. All 3 groups showed segregation among samples of the same classification group. Therefore, based on our results which also compared processing techniques between BF, L-IFs, and P-IFs and their significance, we support Kilvington’s proposal of 7-ketocholesterol as a biomarker for powder IF based on its specific spray drying and canning techniques employed during its confection (A. Kilvington et al., 2019; Risso et al., 2022).

## Data Availability

All data produced in the present study are available upon reasonable request to the authors

## Acknowledgments

This study was funded by the Center for Research Ingredients Safety (CRIS) with the GR100229 grant, and the USDA National Institute of Food and Agriculture, Hatch project MICL02526 to I.G.M.M. The authors are grateful to Kristin Isaacs, Center for Computational Toxicology and Exposure, U.S. Environmental Protection Agency for her help with the SHEDS-HT software package. The authors are grateful to Nama Naseem, Nicole Urrea, Ashley Xu, Rosemary Laurito, Grant Gmitter, and Lisa Zou for their assistance during lipid extraction.

---

**Table 1S:** Food Codes for Baby Foods Used in this Study

| ***Sample ID*** | ***Baby Food*** | ***Brand*** |
| --- | --- | --- |
| BF1 - RTE | Beef and broth/gravy | Beech-Nut |
| BF2 - RTE | Chicken and broth/gravy | Gerber |
| BF3 - RTE | Vegetables and beef | Gerber |
| BF4 - RTE | Vegetables and chicken | Gerber |
| BF5 - RTE | Chicken noodle dinner | Gerber |
| BF6 - RTE | Macaroni, tomato, and cheese | Gerber |
| BF7 - RTE | Turkey, and rice | Gerber |
| BF8 - RTE | Turkey and broth/gravy | Beech-Nut |
| BF9 - RTE | Fruit yogurt | Gerber |
| BF10 - RTE | Chicken with rice | Gerber |
| BF11 - RTE | Vegetables and turkey | Gerber |
| BF12 - RTE | Macaroni and cheese with vegetables | Gerber |
| BF13 - RTE | Pasta pick-ups (cheese ravioli) | Gerber |

**Table 2S:** DOxS Profiling in BF Samples

|  | *BF1* | *BF2* | *BF3* | *BF4* | *BF5* | *BF6* | *BF7* | *BF8* | *BF9* | *BF10* | *BF11* | *BF12* | *BF13* | *Total*<br>*Average*<br>*±STD* |
| --- | --- | --- | --- | --- | --- | --- | --- | --- | --- | --- | --- | --- | --- | --- |
| **Serving Size (g)** | 71.00 | 113.40 | 113.40 | 113.40 | 113.40 | 128.00 | 113.40 | 71.00 | 113.40 | 113.40 | 113.40 | 113.40 | 85.00 | - |
| **Cholesterol**<br>**(mg/100 g fat) ±**<br>**STD** | 1,247.23<br>± 227.45 | 673.84<br>± 98.50 | 58.17 ±<br>8.50 | 319.43<br>± 83.74 | 348.65<br>± 82.26 | 123.24<br>± 29.61 | 263.77<br>± 76.69 | 683.55 ±<br>310.63 | 162.73<br>± 95.67 | 251.02<br>± 57.95 | 313.34<br>± 27.54 | 172.87<br>± 17.84 | 327.40<br>± 87.35 | 380.40 ±<br>307.87 |
| **Cholesterol**<br>**Reference**<br>**(mg/100 g fat)** | 1,000.00 | 833.33 | 250.00 | 500.00 | 400.00 | 7.81 | 400.00 | 875.00 | 333.33 | 333.33 | 285.71 | 333.33 | 500.00 | - |
| **Phytosterols**<br>**(mg/100 g fat) ±STD** |  |  |  |  |  |  |  |  |  |  |  |  |  |  |
| **Brassicasterol** | ND | ND | ND | ND | ND | ND | ND | ND | ND | ND | 19.72 ±<br>3.21 | 29.60 ±<br>3.30 | ND | 24.66 ±<br>4.94 |
| **Campesterol** | ND | ND | 10.48 ±<br>5.14 | 182.76<br>± 53.20 | 136.48<br>± 47.29 | 21.98 ±<br>6.84 | 190.88<br>± 21.15 | ND | ND | 48.06 ±<br>9.69 | ND | 99.33 ±<br>24.73 | 44.86 ±<br>3.55 | 91.85<br>±66.85 |
| **Stigmasterol** | ND | ND | 6.52 ±<br>1.87 | 75.16 ±<br>41.86 | 33.77 ±<br>7.19 | ND | ND | ND | ND | 25.41 ±<br>14.33 | 48.31 ±<br>2.23 | ND | ND | 37.83 ±<br>23.04 |
| **β-Sitosterol** | ND | ND | 47.10 ±<br>13.21 | 498.98<br>± 129.71 | 391.77<br>± 93.78 | 100.31<br>± 19.76 | 421.59<br>± 137.27 | ND | ND | 160.14<br>± 26.56 | 435.62<br>± 41.06 | 236.18<br>± 18.19 | ND | 286.46 ±<br>160.93 |
| **Fucosterol** | ND | ND | ND | ND | ND | ND | ND | ND | ND | ND | ND | ND | 0.046 ±<br>0.014 | 0.046 ±<br>0.00 |
| **Total**<br>**Phytosterols** | ND | ND | 64.1 | 756.9 | 562.02 | 122.29 | 612.47 | 0 | 0 | 233.61 | 503.65 | 365.11 | 44.91 | 251.16 ±<br>264.06 |

\*P-value < 0.001

Table 2S: DOxS Profiling in BF Samples (cont'd)
|  | *BF1* | *BF2* | *BF3* | *BF4* | *BF5* | *BF6* | *BF7* | *BF8* | *BF9* | *BF10* | *BF11* | *BF12* | *BF13* | *Total*<br>*Average*<br>*±STD* |
| --- | --- | --- | --- | --- | --- | --- | --- | --- | --- | --- | --- | --- | --- | --- |
|  | COPs<br>(mg/100 g fat) ±STD |  |  |  |  |  |  |  |  |  |  |  |  |  |
| **7α-OH** | 8.97 ± 5.42 | 2.92 ± 0.73 | 0.10 ± 0.031 | 1.80 ± 0.22 | 2.10 ± 0.50 | 3.73 ± 1.98 | 2.12 ± 0.16 | 5.85 ± 1.82 | 0.68 ± 0.12 | 2.08 ± 0.63 | 0.66 ± 0.092 | 0.41 ± 0.10 | 4.22 ± 3.77 | 2.74 ± 2.40 |
| **7β-OH** | 24.61 ± 15.21 | 2.27 ± 0.64 | 0.065 ± 0.037 | 1.61 ± 0.24 | 1.94 ± 0.37 | 0.66 ± 0.12 | 2.38 ± 0.15 | 4.91 ± 1.12 | 0.59 ± 0.17 | 1.62 ± 0.46 | 0.58 ± 0.020 | 0.40 ± 0.078 | 1.27 ± 0.89 | 3.30 ± 6.27 |
| **5,6α-epoxy** | 0.69 ± 0.36 | 0.044 ± 0.012 | ND | ND | 0.052 ± 0.021 | ND | ND | 0.030 ± 0.019 | ND | 0.027 ± 0.016 | ND | ND | 0.062 ± 0.023 | 0.15 ± 0.24 |
| **5,6β-epoxy** | 1.39 ± 0.81 | 0.033 ± 0.0062 | ND | ND | 0.045 ± 0.027 | ND | ND | 0.073 ± 0.039 | ND | 0.042 ± 0.032 | 1.05 ± 0.12 | ND | 0.10 ± 0.025 | 0.39 ± 0.53 |
| **7-keto** | 16.69 ± 5.45 | 0.84 ± 0.34 | ND | 0.57 ± 0.065 | 0.59 ± 0.18 | 0.67 ± 0.42 | 0.48 ± 0.15 | 1.68 ± 0.61 | 0.24 ± 0.075 | 0.65 ± 0.087 | ND | ND | 1.31 ± 0.83 | 2.37 ± 4.79 |
| **Triol** | 2.63 ± 0.46 | 0.22 ± 0.031 | ND | 0.27 ± 0.086 | 0.31 ± 0.091 | 0.20 ± 0.034 | 0.39 ± 0.056 | 0.27 ± 0.10 | 0.21 ± 0.050 | 0.35 ± 0.041 | 0.12 ± 0.036 | 0.18 ± 0.013 | 0.15 ± 0.03 | 0.44 ± 0.66 |
| **6-keto** | ND | 0.11 ± 0.064 | ND | ND | ND | ND | ND | 0.12 ± 0.018 | ND | ND | ND | 0.10 ± 0.044 | ND | 0.11 ± 0.01 |
| **20α-OH** | ND | ND | 0.65 ± 0.17 | ND | ND | ND | ND | ND | ND | ND | 5.29 ± 0.40 | ND | ND | 2.97 ± 2.32 |
| **22-OH** | ND | ND | 0.07 ± 0.03 | ND | ND | ND | ND | ND | ND | ND | ND | ND | ND | 0.07 ± 0.00 |
| **24-OH** | ND | ND | ND | ND | ND | 0.11 ± 0.06 | 0.13 ± 0.02 | 0.45 ± 0.20 | ND | ND | ND | ND | 0.25 ± 0.19 | 0.24 ± 0.14 |
| **25-OH** | 3.60 ± 1.22 | 0.24 ± 0.055 | ND | 0.16 ± 0.056 | 0.27 ± 0.16 | ND | ND | 0.67 ± 0.16 | ND | 0.27 ± 0.047 | ND | ND | 0.47 ± 0.32 | 0.81 ± 1.15 |
| **Total COPs** | 58.59 ± 27.02 | 6.68 ± 1.80 | 0.88 ± 0.11 | 4.61 ± 0.64 | 5.31 ± 1.22 | 5.37 ± 2.51 | 5.51 ± 0.15 | 14.06 ± 3.95 | 1.71 ± 0.37 | 5.04 ± 1.25 | 7.69 ± 0.42 | 1.08 ± 0.23 | 7.85 ± 6.04 | 9.57 ± 14.53 |

**Table 3S:** Values of Exposure, Dose Intake, and Absorption Provided by SHEDS-HT

|  | Females |  |  | Males |  |  | Total |  |  |
| --- | --- | --- | --- | --- | --- | --- | --- | --- | --- |
|  | *Exposure*<br>*(ug/day)* | *Dose Intake*<br>*(ug/day)* | *Absorption*<br>*(mg/kg-day)* | *Exposure*<br>*(ug/day)* | *Dose Intake*<br>*(ug/day)* | *Absorption*<br>*(mg/kg-day)* | *Exposure*<br>*(ug/day)* | *Dose Intake*<br>*(ug/day)* | *Absorption*<br>*(mg/kg-day)* |
| **Cholesterol** | 77,575.04 | 7.75 | 2.44 | 46,101.42 | 4.44 | 1.40 | 61,391.3 | 6.04 | 1.90 |
| **Desmosterol** | 9,689.138 | 0.99 | 0.38 | 8,286.66 | 0.79 | 0.30 | 8,967.98 | 0.90 | 0.34 |
| **Squalene** | 1,339.386 | 0.13 | 0.13 | 1,576.29 | 0.15 | 0.15 | 1,461.20 | 0.14 | 0.14 |
| **Phytosterols** |  |  |  |  |  |  |  |  |  |
| **Brassicasterol** | 0.087003 | 9.10E-06 | 2.84E-06 | 42.59 | 0.0041 | 0.0013 | 21.94 | 0.0021 | 0.00066 |
| **β-Sitosterol** | 94,659.75 | 9.64 | 0.49 | 57,224.17 | 5.46 | 0.28 | 75410.37 | 7.49 | 0.38 |
| **Campesterol** | 27,609.45 | 2.85 | 0.41 | 20,512.97 | 1.97 | 0.28 | 23,960.44 | 2.40 | 0.34 |
| **Fucoesterol** | 10,530.81 | 1.10 | 0.044 | 7,890.36 | 0.76 | 0.030 | 9,173.092 | 0.93 | 0.037 |
| **Stigmasterol** | 18,091.73 | 1.88 | 0.075 | 13,095.2 | 1.27 | 0.051 | 15,522.51 | 1.56 | 0.063 |
| **Tocopherols** |  |  |  |  |  |  |  |  |  |
| **α-Tocopherol** | 333.66 | 0.034 | 0.0011 | 844.76 | 0.082 | 0.0028 | 596.47 | 0.059 | 0.0020 |
| **β-Tocopherol** | 604.36 | 0.064 | 0.0022 | 866.75 | 0.085 | 0.0029 | 739.28 | 0.075 | 0.0026 |
| **δ-Tocopherol** | 12,780.61 | 1.34 | 0.047 | 8,782.58 | 0.86 | 0.030 | 10,724.83 | 1.09 | 0.038 |
| **γ-Tocopherol** | 2,75.94 | 0.22 | 0.0075 | 1,656.91 | 0.16 | 0.0055 | 1,860.48 | 0.19 | 0.0065 |
| **DOxS** |  |  |  |  |  |  |  |  |  |
| **20-OH** | 0.38 | 4.03E-05 | 1.61E-05 | 1.30 | 0.00013 | 5.03E-05 | 0.86 | 8.42E-05 | 3.37E-05 |
| **22-OH** | 0.30 | 3.12E-05 | 1.25E-05 | 0.21 | 2.04E-05 | 8.16E-06 | 0.25 | 2.56E-05 | 1.03E-05 |
| **24-OH** | 0.32 | 3.29E-05 | 1.32E-05 | 0.31 | 2.98E-05 | 1.19E-05 | 0.31 | 3.13E-05 | 1.25E-05 |
| **25-OH** | 132.46 | 0.014 | 0.0056 | 98.85 | 0.0096 | 0.0038 | 115.17 | 0.012 | 0.0047 |
| **4-cholesten-3,6-dione** | 0 | 0 | 0 | 0.0055 | 5.28E-07 | 2.15E-07 | 0.0028 | 2.71E-07 | 1.1E-07 |
| **6-Keto** | 8.64 | 0.00091 | 0.00037 | 103.38 | 0.010 | 0.0042 | 57.35 | 0.0057 | 0.0023 |
| **7α-OH** | 120.85 | 0.013 | 0.0038 | 207.76 | 0.020 | 0.0060 | 165.54 | 0.017 | 0.0050 |
| **7β-OH** | 127.20 | 0.013 | 0.0056 | 252.86 | 0.024 | 0.010 | 191.81 | 0.019 | 0.0080 |
| **7-Keto** | 36.18 | 0.0038 | 0.00046 | 80.57 | 0.0078 | 0.00094 | 59.01 | 0.0059 | 0.00071 |
| **5,6α-Epoxy** | 4.67 | 0.00049 | 0.00014 | 10.25 | 0.00099 | 0.00028 | 7.54 | 0.00075 | 0.00021 |
| **5,6β-Epoxy** | 732.54 | 0.076 | 0.024 | 1,727.98 | 0.17 | 0.054 | 1,244.39 | 0.12 | 0.040 |
| **Triol** | 45.87 | 0.0048 | 0.00072 | 78.35 | 0.0076 | 0.0011 | 62.57 | 0.0063 | 0.00094 |

**Figure 1S:**
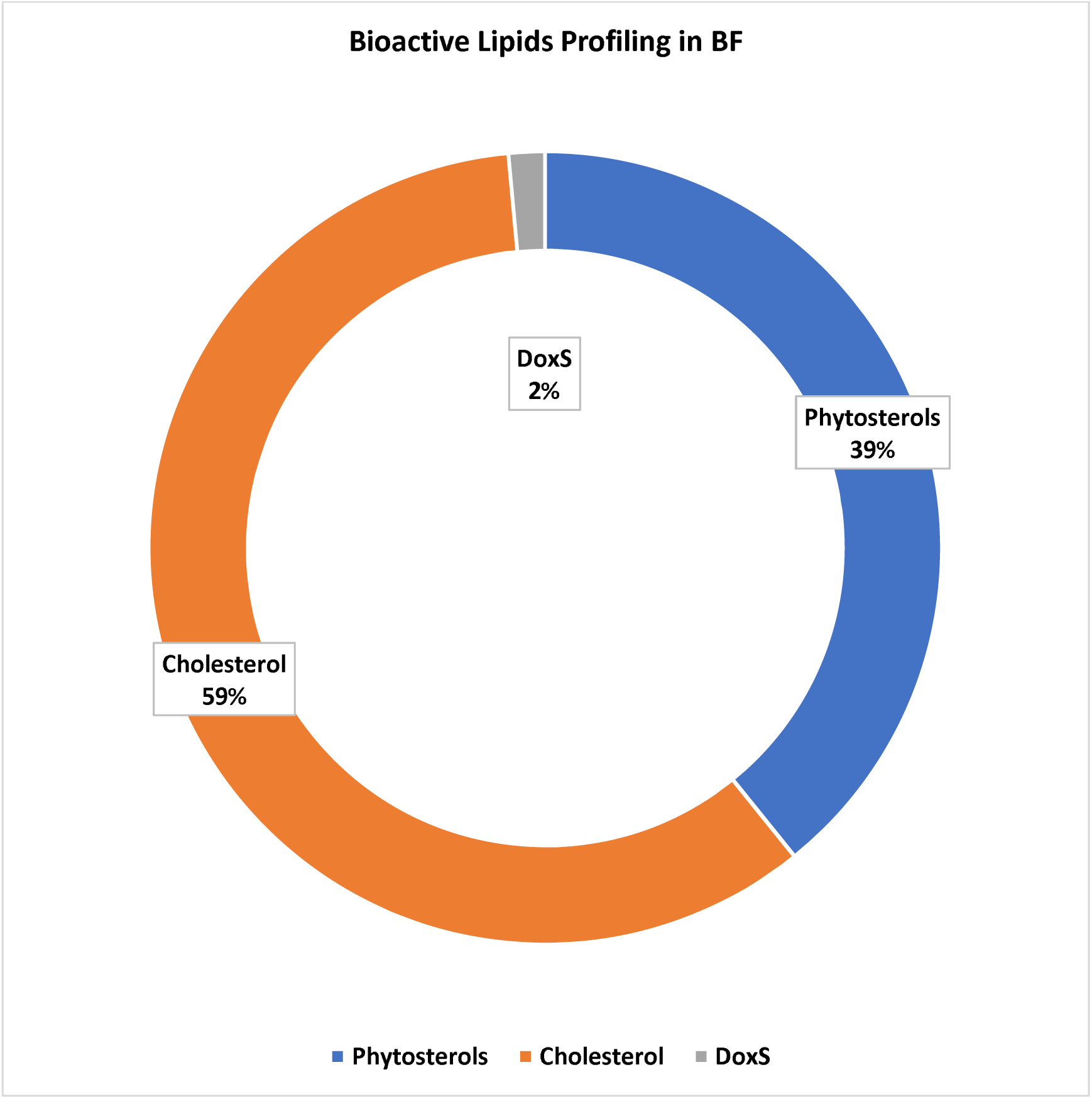
Distribution of Bioactive Lipids Concentrations Among BF

## References

Allen, R. E., & Myers, A. L. (2006). Nutrition in toddlers. Am Fam Physician, 74(9), 1527–1532.

Babawale, E. A., Jones, P. J., Mercer, K. E., Lin, H., Yeruva, L., Bar Yoseph, F., & Rutherfurd, S. M. (2018). Modulating Sterol Concentrations in Infant Formula Influences Cholesterol Absorption and Synthesis in the Neonatal Piglet. Nutrients, 10(12). doi:10.3390/nu10121848

Cais-Sokolińska, D., Walkowiak-Tomczak, D., & Rudzińska, M. (2022). Photosensitized oxidation of cholesterol and altered oxysterol levels in sour cream: Effects of addition of cucumber pickles. J Dairy Sci, 105(6), 4760–4771. doi:10.3168/jds.2022-21856

CDC. (1998). Recommendations to prevent and control iron deficiency in the United States. MMWR

CDC. (2001). Data Table of Infant Weight-for-age Charts. Retrieved from https://www.cdc.gov/growthcharts/html_charts/wtageinf.htm

CDC. (2021a). Maternity Practices in Infant Nutrition and Care (mPINC) Survey. Retrieved from https://www.cdc.gov/breastfeeding/data/mpinc/index.htm

CDC. (2021b). Vitamins & Minerals. Retrieved from https://www.cdc.gov/nutrition/infantandtoddlernutrition/vitamins-minerals/index.html

Centre, T. M. I. (2021). The Metabolomics Innovation Centre. Retrieved from https://www.metabolomicscentre.ca/

ChemSpider. (2021). ChemSpider. Retrieved from http://www.chemspider.com/

Chiu, C.-W. a. K. T.-H. a. C. B.-H. (2018). Improved Analytical Method for Determination of Cholesterol-Oxidation Products in Meat and Animal Fat by QuEChERS Coupled with Gas Chromatography–Mass Spectrometry. Journal of Agricultural and Food Chemistry, 66(13), 3561–3571. doi:10.1021/acs.jafc.8b00250

Chong, J., Soufan, O., Li, C., Caraus, I., Li, S., Bourque, G., Xia, J. (2018). MetaboAnalyst 4.0: towards more transparent and integrative metabolomics analysis. Nucleic Acids Res, 46(W1), W486–W494. doi:10.1093/nar/gky310

Chudy, S., & Teichert, J. (2021). Oxysterols in stored powders as potential health hazards. Scientific Reports, 11(1), 21192. doi:10.1038/s41598-021-00636-5

Claumarchirant, L., Matencio, E., Sanchez-Siles, L. M., Alegría, A., & Lagarda, M. J. (2015). Sterol Composition in Infant Formulas and Estimated Intake. J Agric Food Chem, 63(32), 7245–7251. doi:10.1021/acs.jafc.5b02647

Collective, G. B. (2017). Global Breastfeeding Scorecard 2017, tracking progress for breastfeeding policies and programmes. Paper presented at the UNICEF and World Health Organization, New York and Geneva.

Folch, J., Lees, M., & Sloane Stanley, G. (1957). A simple method for the isolation and purification of total lipides from animal tissues. J biol Chem, 226(1), 497–509.

Fox, M. K., Pac, S., Devaney, B., & Jankowski, L. (2004). Feeding infants and toddlers study: What foods are infants and toddlers eating? J Am Diet Assoc, 104(1 Suppl 1), s22–30. doi:10.1016/j.jada.2003.10.026

Garcia-Llatas, G., Mercatante, D., López-García, G., & Rodríguez-Estrada, M. T. (2021). Oxysterols – how much do we know about food occurrence, dietary intake and absorption? Current Opinion in Food Science. doi:https://doi.org/10.1016/j.cofs.2021.08.001

García-Llatas, G., Cercaci, L., Rodriguez-Estrada, M. T., Lagarda, M. J., Farré, R., & Lercker, G. (2008). Sterol Oxidation in Ready-to-Eat Infant Foods During Storage. Journal of Agricultural and Food Chemistry, 56(2), 469–475. doi:10.1021/jf072475g

García-Llatas, G., & Rodríguez-Estrada, M. T. (2011). Current and new insights on phytosterol oxides in plant sterol-enriched food. Chem Phys Lipids, 164(6), 607–624. doi:10.1016/j.chemphyslip.2011.06.005

Gehring, J., Touvier, M., Baudry, J., Julia, C., Buscail, C., Srour, B., Allès, B. (2020). Consumption of Ultra-Processed Foods by Pesco-Vegetarians, Vegetarians, and Vegans: Associations with Duration and Age at Diet Initiation. The Journal of Nutrition, 151(1), 120–131. doi:10.1093/jn/nxaa196

Gerber. (2021a). Baby Food. In.

Gerber. (2021b). Daily nutrition goals & food groups for Crawlers. Retrieved from https://www.gerber.com/learning-center/crawler-nutrition-basics

Grau, A., Codony, R., Grimpa, S., Baucells, M. D., & Guardiola, F. (2001). Cholesterol oxidation in frozen dark chicken meat: influence of dietary fat source, and alpha-tocopherol and ascorbic acid supplementation. Meat Science, 57(2), 197–208. doi:10.1016/s0309-1740(00)00094-2

Guardiola, F. (2002). Cholesterol and phytosterol oxidation products: analysis, occurrence, and biological effects: The American Oil Chemists Society.

Guardiola, F., Bou, R., Boatella, J., & Codony, R. (2004). Analysis of sterol oxidation products in foods. Journal of Aoac International, 87(2), 441–466.

Hamdan, I. J. A., Claumarchirant, L., Garcia-Llatas, G., Alegría, A., & Lagarda, M. J. (2017). Sterols in infant formulas: validation of a gas chromatographic method. Int J Food Sci Nutr, 68(6), 695–703. doi:10.1080/09637486.2017.1287883

Hamdan, I. J. A., Sanchez-Siles, L. M., Garcia-Llatas, G., & Lagarda, M. J. (2018). Sterols in Infant Formulas: A Bioaccessibility Study. J Agric Food Chem, 66(6), 1377–1385. doi:10.1021/acs.jafc.7b04635

Hamdan, I. J. A., Sanchez-Siles, L. M., Matencio, E., Bermúdez, J. D., Garcia-Llatas, G., & Lagarda, M. J. (2018). Sterols in human milk during lactation: bioaccessibility and estimated intakes. Food Funct, 9(12), 6566–6576. doi:10.1039/c8fo01795f

Hardy, S. C., & Kleinman, R. E. (1994). Fat and cholesterol in the diet of infants and young children: implications for growth, development, and long-term health. J Pediatr, 125(5 Pt 2), S69–77. doi:10.1016/s0022-3476(06)80739-0

Harris, J. L., & Pomeranz, J. L. (2020). Infant formula and toddler milk marketing: opportunities to address harmful practices and improve young children’s diets. Nutr Rev, 78(10), 866–883. doi:10.1093/nutrit/nuz095

Isaacs, K. K., Glen, W. G., Egeghy, P., Goldsmith, M. R., Smith, L., Vallero, D., Ozkaynak, H. (2014). SHEDS-HT: An Integrated Probabilistic Exposure Model for Prioritizing Exposures to Chemicals with Near-Field and Dietary Sources. Environmental Science & Technology, 48(21), 12750–12759. doi:10.1021/es502513w

Jensen, R. G. (1999). Lipids in human milk. Lipids, 34(12), 1243–1271. doi:https://doi.org/10.1007/s11745-999-0477-2

Johnson, R. (1998). Young children and dietary fat intake: how much is too little? J Am Diet Assoc, 98(4), 402. doi:10.1016/S0002-8223(98)00091-1

Khan, M. I., Min, J. S., Lee, S. O., Yim, D. G., Seol, K. H., Lee, M., & Jo, C. (2015). Cooking, storage, and reheating effect on the formation of cholesterol oxidation products in processed meat products. Lipids Health Dis, 14, 89. doi:10.1186/s12944-015-0091-5

Kilvington, A., Barnaba, C., Rajasekaran, S., Laurens Leimanis, M. L., & Medina-Meza, I. G. (2021). Lipid profiling and dietary assessment of infant formulas reveal high intakes of major cholesterol oxidative product (7-ketocholesterol). Food Chem, 354, 129529. doi:10.1016/j.foodchem.2021.129529

Kilvington, A., Barnaba, C., Rajasekaran, S., Laurens, M. L. L., & Gabriela Medina-Meza, I. (2021). Lipid profiling and dietary assessment of infant formulas reveal high intakes of major cholesterol oxidative product (7-ketocholesterol). Food Chemistry, 129529. doi:https://doi.org/10.1016/j.foodchem.2021.129529

Kilvington, A., Maldonado-Pereira, L., Torres-Palacios, C., & Medina-Meza, I. (2019). Phytosterols and their oxidative products in infant formula. Journal of Food Process Engineering, e13151.

Lawrence, R. M., & Lawrence, R. A. (2022). 10 - Normal Growth, Growth Faltering, and Obesity in Breastfed Infants. In R. A. Lawrence & R. M. Lawrence (Eds.), Breastfeeding (Ninth Edition) (pp. 298–325). Philadelphia: Elsevier.

Maldonado-Pereira, L. (2021). Comprehensive Study of the Oxidative Stability of the Most Consumed Ultra-Processed Foods in the USA. (Ph.D.). Michigan State University, Ann Arbor. Retrieved from http://ezproxy.msu.edu/login?url=https://www.proquest.com/dissertations-theses/comprehensive-study-oxidative-stability-most/docview/2536440379/se-2?accountid=12598 Retrieved from http://resolver.ebscohost.com/openurl?ctx_ver=Z39.88-2004&ctx_enc=info:ofi/enc:UTF-8&rfr_id=info:sid/ProQuest+Dissertations+%26+Theses+Global&rft_val_fmt=info:ofi/fmt:kev:mtx:dissertation&rft.genre=dissertations+%26+theses&rft.jtitle=&rft.atitle=&rft.au=Maldonado-Pereira%2C+Lisaura&rft.aulast=Maldonado-Pereira&rft.aufirst=Lisaura&rft.date=2021-01-01&rft.volume=&rft.issue=&rft.spage=&rft.isbn=9798738634932&rft.btitle=&rft.title=Comprehensive+Study+of+the+Oxidative+Stability+of+the+Most+Consumed+Ultra-Processed+Foods+in+the+USA&rft.issn=&rft_id=info:doi/ Dissertations & Theses @ CIC Institutions; ProQuest Dissertations & Theses Global database. (28497722)

Maldonado-Pereira, L., Barnaba, C., De los Campos, G., & Medina-Meza, I. (2021). Evaluation of the Nutritional Quality of Ultra-Processed Foods (Ready to Eat + Fast Food): Fatty Acid Composition. medRxiv, 2021.2004.2016.21255610. doi:10.1101/2021.04.16.21255610

Maldonado-Pereira, L., Schweiss, M., Barnaba, C., & Medina-Meza, I. G. (2018). The role of cholesterol oxidation products in food toxicity. Food and Chemical Toxicology, 118, 908–939. doi:10.1016/j.fct.2018.05.059

Micheletta, F., Natoli, S., Misuraca, M., Sbarigia, E., Diczfalusy, U., & Iuliano, L. (2004). Vitamin E supplementation in patients with carotid atherosclerosis: reversal of altered oxidative stress status in plasma but not in plaque. Arterioscler Thromb Vasc Biol, 24(1), 136–140. doi:10.1161/01.ATV.0000104028.07929.72

Miller, J. L. (2013). Iron deficiency anemia: a common and curable disease. Cold Spring Harbor perspectives in medicine, 3(7), a011866. doi:10.1101/cshperspect.a011866

Musto, M., Faraone, D., Cellini, F., & Musto, E. (2014). Changes of DNA quality and meat physicochemical properties in bovine supraspinatus muscle during microwave heating. Journal of the Science of Food and Agriculture, 94(4), 785–791. doi:10.1002/jsfa.6441

Osada, K., Hoshina, S., Nakamura, S., & Sugano, M. (2000). Cholesterol Oxidation in Meat Products and Its Regulation by Supplementation of Sodium Nitrite and Apple Polyphenol before Processing. Journal of Agricultural and Food Chemistry, 48(9), 3823–3829. doi:10.1021/jf991187k

Pie, J. E. a. S. K. a. S. C. (1991). Cholesterol oxidation in meat products during cooking and frozen storage. Journal of Agricultural and Food Chemistry, 39(2), 250–254. doi:10.1021/jf00002a005

Poli, G., Leoni, V., Biasi, F., Canzoneri, F., Risso, D., & Menta, R. (2022). Oxysterols: From redox bench to industry. Redox Biol, 49, 102220. doi:10.1016/j.redox.2021.102220

Prins, J. R., Schoots, M. H., Wessels, J. I., Campmans-Kuijpers, M. J. E., Navis, G. J., van Goor, H., … Gordijn, S. J. (2022). The influence of the dietary exposome on oxidative stress in pregnancy complications. Molecular Aspects of Medicine, 87, 101098. doi:https://doi.org/10.1016/j.mam.2022.101098

Rant, W., Radzik-Rant, A., Świątek, M., Niżnikowski, R., Szymańska, Ż., Bednarczyk, M., Ślęzak, M. (2019). The effect of aging and muscle type on the quality characteristics and lipid oxidation of lamb meat. Arch Anim Breed, 62(2), 383–391. doi:10.5194/aab-62-383-2019

Risso, D., Leoni, V., Canzoneri, F., Arveda, M., Zivoli, R., Peraino, A., Menta, R. (2022). Presence of cholesterol oxides in milk chocolates and their correlation with milk powder freshness. PLoS One, 17(3), e0264288. doi:10.1371/journal.pone.0264288

Rodríguez-Rodríguez, P., Ramiro-Cortijo, D., Reyes-Hernández, C. G., López de Pablo, A. L., González, M. C., & Arribas, S. M. (2018). Implication of Oxidative Stress in Fetal Programming of Cardiovascular Disease. Frontiers in physiology, 9, 602–602. doi:10.3389/fphys.2018.00602

Rose-Sallin, C. (1995). Quantification of Cholesterol Oxidation Products in Milk Powders Using [2H7]Cholesterol To Monitor Cholesterol Autoxidation Artifacts. Journal of agricultural and food chemistry, 43(4), 935–941.

Sabolová, M., Pohořelá, B., Fišnar, J., Kouřimská, L., Chrpová, D., & Pánek, J. (2017). Formation of oxysterols during thermal processing and frozen storage of cooked minced meat. J Sci Food Agric, 97(15), 5092–5099. doi:10.1002/jsfa.8386

Sander, B. D., Addis, P. B., Park, S. W., & Smith, D. E. (1989). Quantification of Cholesterol Oxidation-Products in a Variety of Foods. Journal of food protection, 52(2), 109–114.

Timby, N., Adamsson, M., Domellöf, E., Grip, T., Hernell, O., Lönnerdal, B., & Domellöf, M. (2021). Neurodevelopment and growth until 6.5 years of infants who consumed a low-energy, low-protein formula supplemented with bovine milk fat globule membranes: a randomized controlled trial. Am J Clin Nutr, 113(3), 586–592. doi:10.1093/ajcn/nqaa354

Timby, N., Domellöf, E., Hernell, O., Lönnerdal, B., & Domellöf, M. (2014). Neurodevelopment, nutrition, and growth until 12 mo of age in infants fed a low-energy, low-protein formula supplemented with bovine milk fat globule membranes: a randomized controlled trial. Am J Clin Nutr, 99(4), 860–868. doi:10.3945/ajcn.113.064295

USDA. (2018). What We Eat in America, NHANES 1999−2018. National Center for Health Statistics: Hyattsville, MD.: Centers for Disease Control and Prevention Retrieved from https://www.cdc.gov/nchs/nhanes/wweia.htm

Wei, S., Wei, Y., Gong, Y., Chen, Y., Cui, J., Li, L., Yi, L. (2022). Metabolomics as a valid analytical technique in environmental exposure research: application and progress. Metabolomics, 18(6), 35. doi:10.1007/s11306-022-01895-7

Willinger, T. (2019). Oxysterols in intestinal immunity and inflammation. J Intern Med, 285(4), 367–380. doi:10.1111/joim.12855

Woollett, L. A., & Heubi, J. E. (2000). Fetal and Neo-natal Cholesterol Metabolism. In Endotext. South Dartmouth, MA: MDText.com, Inc.

Wu, F., Tian, F.-J., & Lin, Y. (2015). Oxidative Stress in Placenta: Health and Diseases. BioMed Research International, 2015, 293271. doi:10.1155/2015/293271

Zmysłowski, A., & Szterk, A. (2019). Oxysterols as a biomarker in diseases. Clin Chim Acta, 491, 103–113. doi:10.1016/j.cca.2019.01.022

Zubin Maslov, P., Hill, J. A., Lüscher, T. F., & Narula, J. (2021). High-sugar feeding and increasing cholesterol levels in infants. Eur Heart J, 42(12), 1132–1135. doi:10.1093/eurheartj/ehaa868

Öhlund, I., Hörnell, A., Lind, T., & Hernell, O. (2008). Dietary fat in infancy should be more focused on quality than on quantity. European Journal of Clinical Nutrition, 62(9), 1058–1064. doi:10.1038/sj.ejcn.1602824

